# The real-world association between male circumcision and risk of HIV infection in sub-Saharan Africa: a household fixed-effects analysis of 279,351 men from 29 countries

**DOI:** 10.1101/2025.05.22.25328138

**Authors:** Caroline A. Bulstra, Xiaochen Dai, Kenneth Ngure, Molly S. Rosenberg, Beatrice M. Wamuti, Jan A.C. Hontelez, Joshua A. Salomon, Katrina F. Ortblad, Till Bärnighausen

## Abstract

**Introduction:** While voluntary medical male circumcision (VMMC) reduces the individual-level risk of HIV acquisition by approximately 60% in randomised-controlled trials, little is known about the ‘real-world’ long-term effect of medical and traditional male circumcision on the cumulative risk of HIV infection. We estimate the association between these for the first time using a quasi-experimental study design—a household fixed-effects analysis—for sub-Saharan Africa, the global region with the largest HIV burden.

**Methods:** We pooled individual-level cross-sectional data from the nationally-representative Demographic and Health Surveys and AIDS Indicator Surveys across all sub-Saharan African countries in which the surveys included data on both male circumcision and HIV status. We estimated the association between male circumcision and HIV status using modified Poisson regression models with household fixed-effects—which control for unobserved and observed confounding shared by men living in the same household—and included additional individual-level controls for demographic characteristics, socio-economic factors, and sexual behaviour.

**Results:** We included individual data from 279,351 male participants in 48 nationally-representative surveys conducted in 29 countries between 2003–2018. The mean survey-level prevalence of male circumcision was 65.9% (median 84.5%, IQR 28.8%–68.1%) and HIV was 5.6% (median 2.5%, IQR 1.2%–10.2%). We estimated that male circumcision was significantly associated with a nearly one-fifth reduction in the cumulative risk of HIV infection (adjusted risk ratio 0.81, 95% CI 0.73–0.89).

**Conclusions:** Male circumcision was associated with a significant reduction in the risk of HIV infection in sub-Saharan Africa over the past two decades. Increased political and financial commitment to VMMC could likely lead to further reductions in HIV prevalence, especially when rolled out as a HIV prevention option in combination with other interventions.

## Introduction

Medical and traditional male circumcision is a common practice in many countries in sub-Saharan Africa. Thus far, little is known about the ‘real-world’ long-term effect of male circumcision on the cumulative risk of HIV-infection. Previous randomised-controlled trials in Uganda, Kenya, and South Africa have established the efficacy of voluntary medical male circumcision (VMMC) [1,2] in reducing HIV incidence [3–6]. Traditional cultural male circumcision practices have been shown to lead to protective effects against HIV, although to a lesser extent as VMMC and with varying levels of efficacy [7–9].

The World Health Organization (WHO) and UNAIDS recommend VMMC for uncircumcised men living in countries with high HIV burden for prevention benefits [2,10]. While the causal efficacy of male circumcision in randomised-controlled trials has been established in terms of reductions in incident HIV infections, the ultimate aim of national VMMC programmes is to reduce population-level HIV incidence, and in turn, population-level HIV prevalence [10,11]. Randomised-controlled trials cannot be used to measure the long-term real-life effects of circumcision on HIV prevalence at a country level. The strongest approaches to causal inference available are thus quasi-experimental methods [12–14]. While mathematical models can be useful to predict the effects of male circumcision on HIV prevalence, these predictions may not materialize in real-world implementation because these models can be reductionist in their representation of reality [15]. Thus, we aim to advance the extant literature on the real-world association between male circumcision and HIV incidence in two ways.

First, we determine the association between male circumcision and the cumulative risk of individual-level HIV infection, which is the individual-level correlate to the population-level measure of HIV prevalence. For the effect estimation we use a household fixed-effects analysis [12,13,16,17]. By including a fixed-effect for each household in our analysis, we control for all unobserved and observed confounding at the level of the household—including household culture, household social position, the distance of a person’s residence to other circumcised men, the distance of a person’s residence to traditional healers and circumcision infrastructures, household income, and expenditures. Second, we quantify the real-life long-term association between male circumcision and the cumulative risk of HIV infection in the global region most affected by HIV during the era of ART scale-up and treatment-as-prevention; the surveys we used cover the period 2003 to 2018. We estimate the association between of all male circumcision types—including medical and traditional circumcision—and the cumulative risk of HIV infection, as men who have already been circumcised as part of a traditional ceremony are no longer eligible for medical circumcision and traditional circumcision confers protective benefits.

Our results will thus provide important insight to policy makers on the real-world association between male circumcision and HIV incidence in sub-Saharan Africa in the era of HIV treatment-as-prevention. Such results, if significant, can boost political and financial commitment to implementation of an efficacious HIV prevention intervention that has not been as vigorously supported as might have been expected based on the randomised-controlled trial findings, leading to male circumcision coverage being below global policy goals [2,18–20].

## Methods

### Setting

Our analyses focused on countries across sub-Saharan Africa. In 2021, HIV prevalence among people ages 15-49 years was 6.7% in Eastern and Southern Africa and 1.4% in Western and Central Africa [21]. Male circumcision prevalence was >95% in settings where traditional circumcision is common practice—e.g., Western and Central Africa, but generally <40% in settings where it is not common practice—e.g., Eastern and Southern Africa [22]. From 2008- 2012, national VMMC programmes were initiated and scaled-up in 15 countries including: eSwatini, Kenya, Malawi, South Africa, and Zambia starting in 2008; Ethiopia, Mozambique, Namibia, Rwanda, Tanzania and Zimbabwe starting in 2009; Uganda and Rwanda starting in 2010; Lesotho starting in 2012; and South Sudan starting in 2017 [20,21].

### Survey data

We used individual-level, cross-sectional Demographic and Health Survey (DHS) and AIDS Indicator Survey (AIS) data for our analyses. The DHSs are nationally-representative household surveys that include a wide range of socio-economic, (sexual) behavioural, and epidemiological variables. The AISs are nationally-representative household surveys that were developed in addition to the DHS to measure additional indicators for effective monitoring of national HIV programmes. In both DHS and AIS, around 350 sample locations or ‘clusters’ are randomly sampled throughout the country. Within each cluster, around 25 households are sampled and all willing residents in the household are independently interviewed. Self-reported male circumcision was included in most men’s questionnaires in both DHS and AIS surveys.

Most DHS and AIS in sub-Saharan Africa also include HIV status data. HIV status is determined via voluntary, blood-based HIV testing that uses an enzyme-linked immunosorbent assay (ELISA). More details on survey protocols and questionnaires can be found on the DHS website (https://dhsprogram.com/).

We included all DHSs and AISs that (i) were conducted between 2000 and 2020 in sub-Saharan African countries and (ii) contained both male circumcision and HIV status data in the same survey in our analysis. This resulted in a dataset that comprised 48 surveys from 29 countries. An overview of the available surveys by country, year, and survey type are provided in **Table 1** and **Supplementary** Figure 1. A flow chart of the data selection is provided in Supplementary Figure 2.

**Table 1.** Survey characteristics by African region. Unweighted HIV and circumcision prevalence estimates.

| African region, country | Survey year(s) | Sample size (N) | Survey type | HIV-positive men (HIV prevalence) | Circumcised men (circumcision prevalence) |
| --- | --- | --- | --- | --- | --- |
| **Central Africa** |  |  |  |  |  |
| Burundi | 2010 | 4,078 | DHS | 56 (1.4%) | 1,450 (35.6%) |
|  | 2016-17 | 7,377 | DHS | 53 (0.7%) | 3,191 (43.3%) |
| Chad | 2014-15 | 4,925 | DHS | 61 (1.2%) | 4,748 (96.8%) |
| Congo DR | 2007 | 4,305 | DHS | 43 (1.0%) | 4,167 (97.1%) |
| Gabon | 2012 | 5,502 | DHS | 168 (3.1%) | 5,460 (99.3%) |
| **Eastern Africa** |  |  |  |  |  |
| Ethiopia | 2011* | 13,015 | DHS | 182 (1.4%) | 11,987 (92.3%) |
|  | 2016* | 11,327 | DHS | 115 (1.0%) | 10,443 (92.5%) |
| Kenya | 2008-09* | 3,095 | DHS | 154 (5.0%) | 2,597 (83.9%) |
| Rwanda | 2005 | 4,728 | DHS | 115 (2.4%) | 929 (19.7%) |
|  | 2010* | 6,296 | DHS | 154 (2.4%) | 853 (13.6%) |
|  | 2014-15* | 6,191 | DHS | 161 (2.6%) | 1,803 (29.1%) |
| Tanzania | 2007-08 | 6,333 | AIS | 199 (3.1%) | 4,822 (76.4%) |
| Uganda | 2011* | 9,920 | AIS | 595 (6.0%) | 2,753 (27.8%) |
| **Southern Africa** |  |  |  |  |  |
| Angola | 2015-16 | 5,150 | DHS | 70 (1.4%) | 4,953 (96.6%) |
| Eswatini | 2006-07 | 3,602 | DHS | 704 (19.5%) | 298 (8.3%) |
| Lesotho | 2004 | 2,797 | DHS | 415 (18.6%) | 1,433 (51.4%) |
|  | 2009 | 3,075 | DHS | 543 (17.7%) | 1,724 (56.1%) |
|  | 2014* | 2,775 | DHS | 527 (19.0%) | 2,066 (74.5%) |
| Malawi | 2010* | 6,512 | DHS | 530 (8.1%) | 1,203 (18.5%) |
|  | 2015-16* | 6,855 | DHS | 477 (7.2%) | 1,738 (25.4%) |
| Mozambique | 2009* | 4,404 | AIS | 442 (10.0%) | 2,259 (51.4%) |
|  | 2015* | 4,436 | AIS | 480 (10.9%) | 2,667 (60.2%) |
| Namibia | 2013* | 3,874 | DHS | 419 (10.8%) | 995 (25.8%) |
| South Africa | 2016* | 2,136 | DHS | 310 (14.5%) | 1,170 (54.9%) |
| Zambia | 2007 | 5,161 | DHS | 649 (12.6%) | 805 (15.6%) |
|  | 2013-14* | 13,574 | DHS | 1,573 (11.6%) | 3,316 (24.5%) |
|  | 2018* | 11,547 | DHS | 925 (8.0%) | 3,642 (31.6%) |
| Zimbabwe | 2005 | 5,566 | DHS | 782 (14.1%) | 593 (10.7%) |
|  | 2010-11* | 7,480 | DHS | 811 (13.4%) | 696 (9.4%) |
|  | 2015* | 7,420 | DHS | 889 (12.0%) | 1,205 (16.3%) |
| **Western Africa** |  |  |  |  |  |
| Burkina Faso | 2003 | 3,605 | DHS | 46 (1.4%) | 3,069 (85.1%) |
|  | 2010 | 7,039 | DHS | 60 (0.9%) | 6,119 (87.0%) |
| Cameroon | 2004 | 5,044 | DHS | 215 (3.1%) | 4,782 (94.9%) |
|  | 2011 | 6,948 | DHS | 759 (3.9%) | 6,629 (95.5%) |
| Cote d'Ivoire | 2005 | 19,650 | AIS | 127 (2.9%) | 18,798 (96.2%) |
|  | 2011-12 | 4,352 | DHS | 43 (1.3%) | 4,197 (96.5%) |
| Ghana | 2003 | 5,015 | DHS | 45 (1.1%) | 4,648 (92.7%) |
|  | 2014 | 4,161 | DHS | 35 (1.2%) | 3,860 (92.8%) |
| Guinea | 2005 | 2,930 | DHS | 56 (1.5%) | 2,896 (98.9%) |
| Liberia | 2007 | 5,207 | DHS | 58 (1.1%) | 5,093 (98.7%) |
|  | 2013 | 3,805 | DHS | 45 (1.2%) | 3,767 (99.2%) |
| Mali | 2006 | 3,886 | DHS | 32 (1.0%) | 3,789 (97.6%) |
|  | 2012-13 | 3,751 | DHS | 31 (0.8%) | 3,680 (98.3%) |
| Niger | 2006 | 3,232 | DHS | 32 (1.0%) | 3,215 (99.5%) |
| Senegal | 2005 | 3,251 | DHS | 13 (0.4%) | 3,185 (98.1%) |
| Sierra Leone | 2008 | 3,009 | DHS | 32 (1.1%) | 2,900 (97.9%) |
|  | 2013 | 6,735 | DHS | 81 (1.2%) | 6,694 (99.6%) |
| Togo | 2013-14 | 4,365 | DHS | 72 (1.6%) | 4,240 (97.1%) |
| **Sub-Saharan Africa (all data)** |  |  |  |  |  |
| Survey-level mean, median (IQR) | N/A | 344,832 | N/A | 15,652 (4.7%) | 181,696 (64.7%) |

\* Data collected in country and year when national voluntary medical male circumcision (VMMC) programmes in place.

### Variables

Our main variables of interest were male circumcision (self-reported as circumcised or not circumcised, not restricted by type of circumcision) and HIV status (based on ELISA blood test results; HIV-positive or HIV-negative). We extracted the following additional variables from the DHS and AIS data: sociodemographic variables (age, educational attainment, marital status, and household wealth); sexual behaviours (number of lifetime sex partners, number of sex partners in the past 12 months, and diagnosed sexual transmitted infection [STI] or signs of a STI—e.g., genital sore/ulcer or genital discharge—in the past 12 months); and circumcision variables, including circumcision status and type of circumcision (medical or traditional).

### Statistical analyses

We used a household fixed-effects analysis [12,13,23,24] to estimate the association between circumcision and the cumulative risk of HIV infection (among males aged 15 years or older) up to the average age at which men participated in the surveys. Since household fixed-effects control for unobserved confounding, they are often classified as quasi-experiments [13]. While household fixed-effects control for all confounding at the household level, including unobserved confounding, they do not control for confounding at the individual level. Thus, we included the following potential individual-level variables in our analysis to control for within household individual-level confounding: socio-economic variables (age, highest completed education, marital status) and sexual behaviour (number of lifetime sex partners and having had an STI or symptoms of an STI in the past 12 months). To estimate adjusted risk ratios (aRR), we used modified Poisson models with HIV status as our dependent variable and household fixed-effects, controlling for the individual variables described above. We chose to estimate risk ratios over odds ratios (using other binary models) because they are easier for policy makers to understand and interpret [25].

### Sensitivity analyses

We ran different sensitivity analyses to verify the robustness of our estimates: (1) an unadjusted model (does not control for any additional individual-level variables), (2) a model adjusted only for age, and (3) a model adjusted for age and educational attainment, (4) a model adjusted for age, educational level, and marital status, and (5) the main model applied to the subsample of men who live in the same household but differ in male circumcision status (discordant men).

### Sub-group analyses

We performed various sub-group analyses to test the robustness of our outcomes and estimate potential heterogeneity in estimated associations across different sub-groups of interest. First, we conducted separate analyses for men from Central, Eastern, Southern, and Western Africa (regions according to the African Union [26]) to understand how associations vary in relation to the geographical location of the participants in our study (i.e., Western and Central Africa with higher proportions of traditional circumcision versus Eastern and Southern Africa with higher proportions of VMMC). Second, we conducted separate analyses for younger (aged 15- 34 years) and older (aged 35-54 years) males to explore possible effect modification by age. Third, we performed separate analyses for surveys conducted at times when national VMMC programmes were in place (i.e., launched at least one year before collection of the survey data) versus when no national VMMC programmes were in place to understand the association between circumcision and HIV infection during times of VMMC access. We obtained data on the start dates of national VMMC programmes from the latest available WHO and UNAIDS VMMC progress report [27], an overview is provided in **Supplementary Table 1**. The priority region for VMMC is shown in **Supplementary** Figure 3. Finally, we conducted separate analyses by family relatedness, i.e., for brothers living in the same household, to explore possible biological heterogeneities in the effect of male circumcision on HIV status.

Data management and analyses were done using R version 3.6.2 and Stata version 15.0, maps were created using ArcGIS version 2.3.0.

### Ethical approval

The study was conducted using DHSs and AISs surveys from the USAID DHS programme, which are available upon request. No ethical approval or additional consent was required. All individual-level data was anonymized before distribution. Details of the ethical review process of DHS are available on the programme’s website https://www.dhsprogram.com/What-We-Do/Protecting-the-Privacy-of-DHS-Survey-Respondents.cfm.

## Results

We obtained individual-level data from 29 countries and 48 surveys between 2003 and 2018 (**Table 1**). In the sample of complete-case analysis, 344,832 men from 241,447 different households were included. The number of men per survey ranged from 2,136 men (South Africa, 2016) to 19,650 men (Cote d’Ivoire, 2015-16). For 279,351 men (81.0% of the total sample), both circumcision and HIV status were available. These men resided in 195,803 different households; 94,609 men (33.9%) came from households where two or more men were included in the data subset. Survey-level mean (unweighted) HIV prevalence levels varied by African region, ranging from 0.5% to 3.1% in Central Africa, 0.5% to 3.9% in Western Africa, 1.0% to 6.0% in Eastern Africa, and 1.4% to 19.5% in Southern Africa. Unweighted self-reported mean male circumcision prevalence ranged from 8.9% to 96.6% in Southern Africa, 13.6% to 92.5% in Eastern Africa, 35.6% to 99.3% in Central Africa, and 85.1% to 99.6% in Western Africa.

The socio-demographic characteristics for the men included in the sample—broken down by men living with HIV (4.7%, n=15,952) and circumcised men (64.7%, n=183,460)—are presented in **Table 2**. HIV prevalence was lower in circumcised (3.5%, n=6,257) versus uncircumcised (8.6%, n=8,440) men. In the sample, 38.5% (n=132,724) of men were between 15-24 years of age and HIV prevalence was lowest among this age group (1.6%) and highest among men 35-44 years of age (8.2%), followed by men 45-54 years (7.2%). There was little variation in circumcision prevalence by age group.

**Table 2.** Characteristics of our data sample.

|  | All included men<br>N (% of total) | Number of men<br>living with HIV<br>(HIV<br>prevalence, %)* | Number of<br>circumcised men<br>(male<br>circumcision<br>prevalence, %)* |
| --- | --- | --- | --- |
| **Sample size** | 344,832 | 15,952 (4.7) | 183,460 (64.7) |
| **HIV status** |  |  |  |
| Positive | 15,952 (4.7) | N/A | 6,257 (42.6) |
| Negative | 322,794 (95.3) | N/A | 175,004 (66.1) |
| **Male circumcision status** |  |  |  |
| Circumcised | 183,460 (64.7) | 6,257 (3.4) | N/A |
| Not circumcised | 100,088 (35.3) | 8,440 (8.4) | N/A |
| **Sociodemographic characteristics** |  |  |  |
| **Age (per 10-year age group)** |  |  |  |
| 15-24 | 132,724 (38.5) | 2,045 (1.6) | 67,435 (61.9) |
| 25-34 | 91,326 (26.5) | 4,963 (5.5) | 49,928 (66.4) |
| 35-44 | 67,968 (19.7) | 5,468 (8.2) | 37,309 (67.1) |
| 45-54 | 42,206 (12.2) | 2,976 (7.2) | 23,105 (66.0) |
| 55+ | 10,608 (3.1) | 500 (4.7) | 5,683 (65.0) |
| **Highest education** |  |  |  |
| No education | 75,403 (21.9) | 1,859 (2.5) | 48,779 (82.0) |
| Primary | 129,201 (37.5) | 6,648 (5.2) | 59,434 (55.2) |
| Secondary | 120,958 (35.1) | 6,398 (5.4) | 63,559 (63.7) |
| Post-secondary | 19,245 (5.6) | 1,046 (5.6) | 11,680 (71.0) |
| **Socioeconomic status** |  |  |  |
| 1 'Poorest' | 66,980 (19.4) | 2,422 (3.7) | 35,093 (64.9) |
| 2 | 66,485 (19.3) | 2,698 (4.1) | 34,643 (63.9) |
| 3 | 65,708 (19.1) | 2,970 (4.6) | 33,607 (62.2) |
| 4 | 67,275 (19.5) | 3,707 (5.6) | 34,781 (62.7) |
| 5 'Wealthiest' | 78,381 (22.7) | 4,155 (5.5) | 45,335 (69.0) |
| **Marital status** |  |  |  |
| Never married | 141,046 (41.6) | 2,917 (2.1) | 73,539 (62.8) |
| Currently married | 184,183 (54.3) | 10,880 (6.0) | 101,967 (66.0) |
| Previously married | 13,941 (4.1) | 1,885 (13.8) | 7,953 (66.2) |
| **Family relatedness** |  |  |  |
| Brothers (living in the same household) | 73,010 (22.5) | 1,865 (2.6) | 35,633 (48.8) |
| Other | 250,959 (77.5) | 13,262 (5.3) | 127,231 (50.7) |
| **(Sexual) behavioural characteristics** |  |  |  |
| **Lifetime sex partners** |  |  |  |
| None | 97,150 (29.2) | 1,973 (2.1) | 50,756 (65.6) |
| 1-2 | 89,459 (26.9) | 2,375 (2.7) | 44,379 (59.8) |
| 3-6 | 88,331 (26.5) | 5,809 (6.6) | 45,619 (61.5) |
| 7+ | 58,127 (17.5) | 4,910 (8.5) | 34,485 (72.5) |
| **Had STI during past 12 months** |  |  |  |
| Yes | 18,541 (5.4) | 1,819 (9.9) | 10,010 (63.8) |
| No | 325,962 (94.6) | 14,122 (4.4) | 173,321 (64.8) |
| **Ever engaged in transactional sex** |  |  |  |
| Yes | 19,794 (11.1) | 1,844 (9.5) | 9,392 (58.8) |
| No | 159,146 (88.9) | 7,213 (4.6) | 84,905 (64.3) |
| **Survey characteristics** |  |  |  |
| **Region** |  |  |  |
| Central Africa | 41,388 (12.0) | 599 (1.5) | 20,125 (73.8) |
| Eastern Africa | 83,661 (24.3) | 2,401 (3.0) | 39,247 (61.0) |
| Southern Africa | 98,768 (28.6) | 10,789 (11.2) | 32,527 (33.8) |
| Western Africa | 121,015 (35.1) | 2,163 (1.8) | 91,561 (95.7) |
| **Time of survey** |  |  |  |
| Between 2000-2004 | 40,308 (11.7) | 1,451 (3.9) | 28,853 (88.1) |
| Between 2005-2009 | 106,046 (30.8) | 4,965 (4.7) | 71,250 (76.3) |
| Between 2010-2014 | 136,072 (39.5) | 6,111 (4.5) | 60,358 (57.3) |
| Between 2015-2019 | 62,406 (18.1) | 3,425 (5.5) | 22,999 (44.1) |
| **National VMMC programmes in place** |  |  |  |
| Yes | 127,751 (39.4) | 8,963 (7.0) | 51,240 (40.1) |
| No | 196,218 (60.6) | 6,164 (3.1) | 111,624 (56.9) |

\*Note that the percentages between brackets represent the HIV prevalence and male circumcision prevalence for each category.

The African regions and included countries are shown in **Supplementary** Figure 3. The prevalence of circumcision among men in sub-Saharan Africa is shown in **Figure 1: panels A, B and C**. Male circumcision varied widely across countries and across the African regions. In Western and Central Africa, circumcision prevalence was consistently high: over 85% of men, or in some countries over 98% of men, were circumcised. In Eastern and Southern Africa, larger variability in circumcision prevalence was observed: with prevalence levels of below 30% or 60% in the majority of the Southern African countries (e.g., in Malawi, Zambia, Zimbabwe, Namibia, South Africa, and Lesotho), but circumcision prevalence levels of above 60% in Ethiopia and Kenya. The prevalence of HIV among men in sub-Saharan Africa is shown in **Figure 1: panels D, E and F**. HIV prevalence was highest (above 15%) in eSwatini and Lesotho. Namibia, South Africa, Zimbabwe, Malawi, and Mozambique had an HIV prevalence of between 10% and 15%. HIV prevalence was generally low below 1% or below 2%, in countries throughout Western and Central Africa.

**Figure 1.**
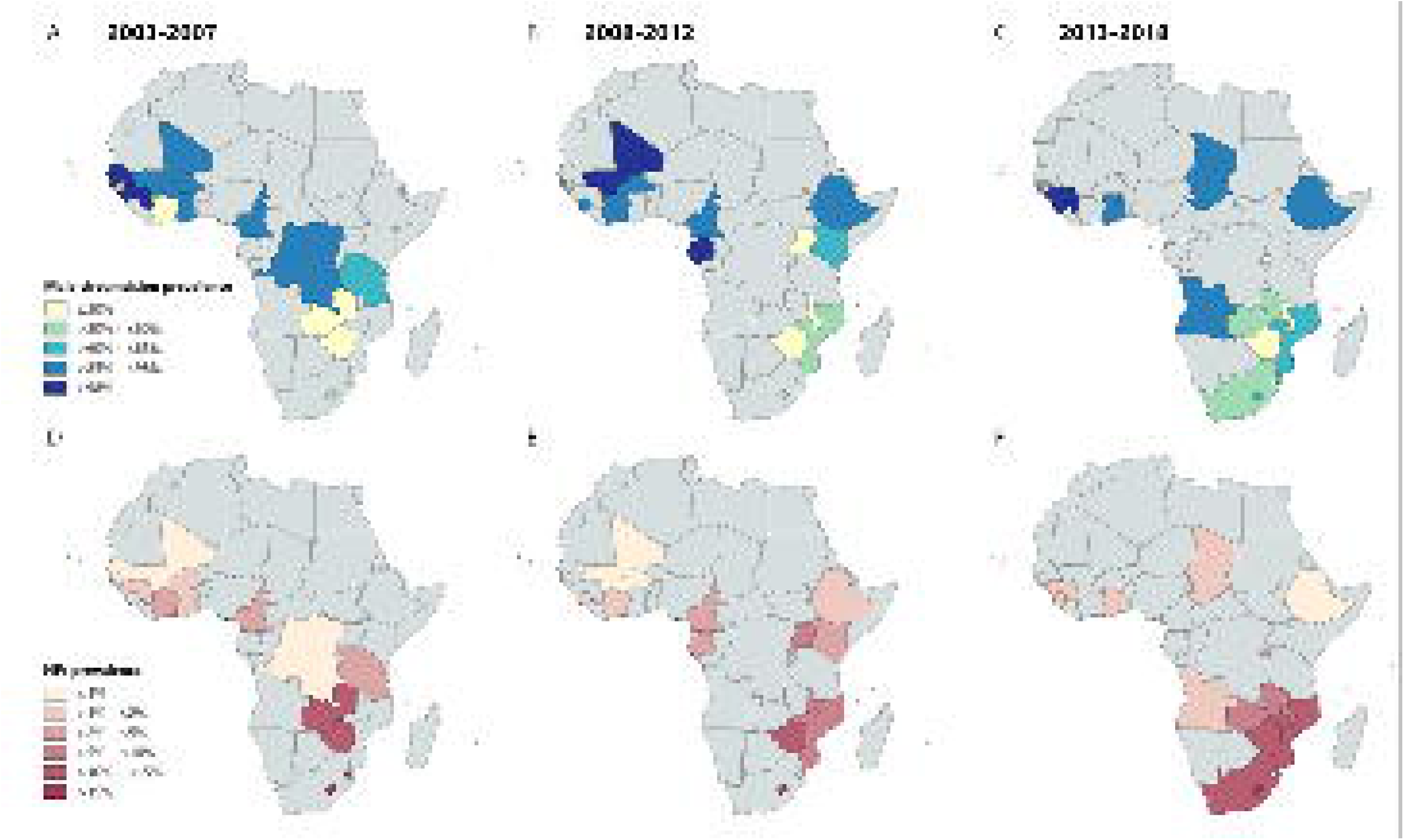
Maps show the national-level male circumcision prevalence from the surveys conducted between 2003-2007 (panel A), 2008-2012 (panel B), and 2013-2018 (panel C) and national-level HIV prevalence levels in men from the surveys conducted between 2003-2007 (panel D), 2008-2012 (panel E), and 2013-2018 (panel F). Countries without surveys for the representative time period are coloured grey. Data available via https://www.dhsprogram.com/.

Figure 1 shows geographical maps visualising national-level male circumcision prevalence and HIV prevalence from the DHSs and AISs in our sample. If multiple surveys were available for a country, the mean weighted prevalence levels were displayed.

In our main analysis, the aRR for living with HIV was 0.81 (95% confidence interval [CI] 0.73– 0.89) among circumcised men compared to uncircumcised men in sub-Saharan Africa, which corresponds to a 19% (95% CI 11%–27%) real-world reduction in HIV risk at the population level (**Table 3**). These findings were confirmed in our four sensitivity analyses, which resulted in similar outcomes (**Supplementary Table 2**) as well as in the main model based on the subsample of men living in the same household but with discordant circumcision status (**Supplementary Table 3**).

**Table 3.** The association between male circumcision and HIV infection among (males aged 15 years and older) in sub-Saharan Africa.

|  | **Main model – adjusted for age, educational level, marital status and sexual behaviour** |  |
| --- | --- | --- |
| **Variable** | **aRR [95% CI]** | **p-value** |
| **Male circumcision status** |  |  |
| Circumcised | 0.81 [0.73–0.89] | <0.001 |
| Not circumcised | 1 (ref) | - |
| **Age** |  |  |
| 15-24 | 1 (ref) | - |
| 25-34 | 2.30 [2.06–2.57] | <0.001 |
| 35-44 | 2.92 [2.57–3.32] | <0.001 |
| 45-54 | 3.10 [2.70–3.57] | <0.001 |
| 55+ | 2.15 [1.71–2.71] | <0.001 |
| **Highest education** |  |  |
| No education | 1.06 [0.94–1.21] | 0.309 |
| Primary | 1 (ref) | - |
| Secondary | 1.00 [0.92–1.09] | 0.941 |
| Post-secondary | 1.00 [0.85–1.17] | 0.961 |
| **Marital status** |  |  |
| Never married | 1 (ref) | - |
| Currently married | 1.51 [1.36–1.68] | <0.001 |
| Previously married | 2.01 [1.77–2.28] | <0.001 |
| **Lifetime sex partners** |  |  |
| None | 0.99 [0.86–1.14] | 0.902 |
| 1-2 | 1 (ref) | - |
| 3-6 | 1.20 [1.08–1.32] | <0.001 |
| 7+ | 1.30 [1.16–1.45] | <0.001 |
| **Had STI during past 12 months** |  |  |
| Yes | 1 (ref) | - |
| No | 1.45 [1.30–1.61] | <0.001 |
| **Model summary** |  |  |
| N (observations) | 18,836 |  |
| Pseudo R<sup>2</sup> | 0.239 |  |

Adjusted risk ratios (aRR) are shown with 95% confidence intervals (CIs). The analysis includes household-level fixed-effects. Schooling levels are defined as the highest level of education attained. Previously married includes being divorced, separated, or widowed. The number of observations (N) includes men from all households where at least two circumcision discordant men (*i.e.* at least one man circumcised and one uncircumcised).

The estimated HIV risk reduction for circumcised men varied somewhat across regions, although smaller stratified samples prohibit precise comparison: Eastern Africa (aRR 0.63, 95% CI 0.46–0.85, p = 0.002), Southern Africa (aRR 0.81, 95% CI 0.72–0.91, p <0.001), Central Africa (aRR 0.88, 95% CI 0.43–1.82, p = 0.727), Western Africa (aRR 0.84, 95% CI 0.60–1.16, p = 0.285) (Figure 2). Outcomes from the other three sub-group analyses (age, prevalence of a national VMMC programs, and brothers) are shown in **Supplementary Table 4**. Male circumcision decreased the cumulative risk of HIV infection for both younger (≤34 years) and older men (≥35 years), but was significant in younger men (aRR 0.79, 95% CI 0.69–0.91, p <0.001) and not in older men (aRR 0.82, 95% CI 0.63–1.06, p = 0.130). Male circumcision significantly reduced the cumulative risk of HIV infection in country-years with national VMMC programmes in place (aRR 0.75, 95% CI 0.66–0.85, p <0.001), but was not significantly associated with risk of HIV infection in country-years without national VMMC programmes (aRR 0.98, 95% CI 0.83–1.15, p = 0.768). In the sub-group analysis of brothers, the association between circumcision and HIV infection was similar to the estimate in the main analysis (aRR 0.78, 95% CI 0.66–0.91, p = 0.002).

**Figure 2.**
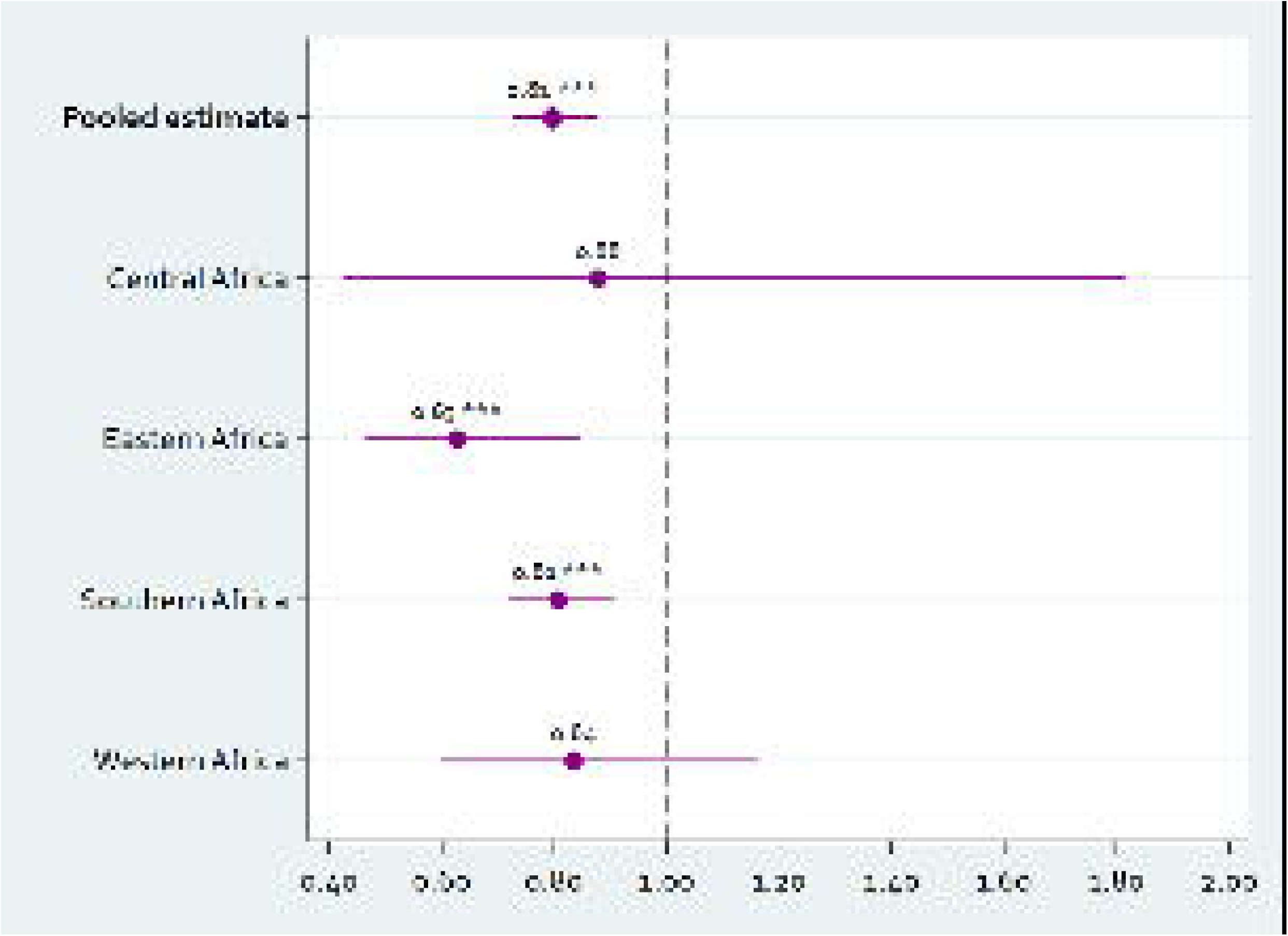
Population-level association between male circumcision and HIV infection (males aged 15 years or older) in the pooled sub-Saharan Africa sample and by African region. The figure shows the adjusted risk ratios (aRRs) and 95% confidence intervals (CIs) from modified Poisson regression models with household fixed-effects. All models were adjusted for age (by 10-year age group), educational level, and marital status. The RRs represent the effects of male circumcision on the cumulative risk of HIV infection. The full regression output of the pooled dataset and sub-group analyses by African region are presented in **Table 3** and Supplementary Table 4 respectively. Data available via https://dhsprogram.com/.

We checked the representativeness of our sample by comparing men from circumcision status discordant households with men in the overall population (**Supplementary Table 5**).

## Discussion

In this quasi-experimental analysis using nationally-representative data from 48 surveys in 29 sub-Saharan African countries, male circumcision was associated with a cumulative one-firth risk reduction in HIV infection among males aged 15 years or older. Current male circumcision coverage in many Eastern and Soutern African countries remains low. Our findings thus further emphasize the potential of medical male circumcision for reducing the long-term burden of HIV in the world regions where two-thirds of people living with HIV reside [28]. Importantly, the large our study established significant real-life, long-term associations between male circumcision and HIV infection during a period of ART scale-up to very high levels of coverage levels in sub-Saharan African (2003-2018). Thus, male circumcision has strong complementary effects to the population HIV prevention benefits of ART scale up and remains an important preventive intervention in the context of treatment-as-prevention.

Our analyses showed lower HIV protective effects of male circumcision for Western and Central African countries and, generally, for countries without national VMMC programmes in place (at time of the survey conducted). This may be attributed to a discrepancy in risk reduction from traditional male circumcision as compared to VMMC and higher protective effects in high HIV incidence settings. Prospective cohort studies from rural Uganda [29] and rural Kenya [7,30], showed traditional male circumcision to be associated with an around 2-fold decrease in HIV acquisition, relative to the 3-fold decrease with VMMC found in the randomised-controlled trials. However, similar studies from Central, Southern and Western Africa are lacking. Experimental and observational studies on the association between traditional male circumcision compared to VMMC and HIV incidence in different contexts outside of Eastern Africa should point out whether there is heterogeneity in association between studies from the different African regions. Although circumcision modality (traditional or medical) was available for some of the men included in our pooled data sample, missingness was very high (69.1%) and hence a household fixed-effect analysis is not reliable due to the low number households with men who showed circumcision status discordance when selecting for traditional circumcision. Therefore, we cannot make definite conclusion on the association between VMMC programmes and HIV prevalence. While there might be a significant association between male circumcision and reduced HIV incidence, this might not (yet) be reflected in HIV prevalence levels, due to the relatively recent scale-up of VMMC programmes and young target population [31]. One of the major advantages of male circumcision, compared to other HIV prevention interventions, is that its effectiveness does not rely on repeated and consistent behaviours [32].

We confirm that male circumcision should be recognised as an important means to reduce the risk of HIV acquisition, thereby indirectly protecting women and children from HIV infection [33]. especially when rolled out as a HIV prevention option in combination with other interventions.

Based on our findings, VMMC could be promoted as a HIV prevention option, alongside other effective behavioural and biomedical prevention interventions (e.g., treatment-as-prevention [TasP] and pre-exposure prophylaxis [PrEP]). Overall, medical male circumcision should primarily be rolled-out in communities with high HIV incidence and prevalence levels where male circumcision is not traditionally done, and could be targeted at men at higher risk of acquiring HIV such, as truck drivers and the clients of sex workers [34].

This study has a number of limitations. First, multivariable regression analyses with household fixed-effects can only provide hints on causal inference, but not claim causality with the same strength as randomised-controlled trials. Using this model, we control for all observed and unobserved household-level confounding, but there is limited control of individual-level confounding (i.e. unobserved individual-level confounding is not accounted for, but observed individual-level confounding is through the included model covariates) [13,23]. Although fixed-effect models have been widely applied in econometrics and economics [24], the use in global health is relatively new, and findings should be interpreted in the light of existing evidence from traditional study designs. We do believe that the use of household fixed-effect regression models on large (pooled) cross-sectional datasets forms a very promising complement to randomised-controlled trials (which are causally strong, but often conducted at small geographical scales and with weak external validity) and observational studies (which are often conducted at larger geographical scales, but associative rather than causal). Second, the sample represents a random selection of males aged 15 years with at least one other man in the same household in this age range with discordant circumcision status. Selection of households with discordant male circumcision status could have biased our associations compared to the unobserved circumcision association in the entire population. We checked the representativeness of our sample for all males aged 15 years or older in the surveys by comparing observed characteristics of the men included in our analyses with men in the overall population (**Supplementary Table 5**). As expected, due to likely higher rates in medical circumcision (as compared to traditional circumcision) in discordant households, men from discordant households were more often younger, higher educated, and wealthier, but discrepancies were modest.

## Conclusions

In conclusion, our findings indicate that male circumcision is associated with a reduced the risk of HIV infection among men in sub-Saharan Africa in the ‘real-world’ context of ART scale-up and the era of treatment-as-prevention. These findings underpin the importance of scaling up VMMC, alongside other HIV prevention interventions, in HIV control and elimination programmes. Given overall low levels of circumcision coverage in many countries in sub-Saharan Africa, the potential for impact of circumcision on HIV burdens remains large.

## Competing interests

All authors are salaried employees of the institutions to which they are affiliated in the header. No specific funding was granted to support this study. All authors declare no competing interests.

### Authors’ contributions

CAB and TB initiated and conceptualised the study. CAB, XD, KFO and TB wrote the first draft of the manuscript and verified the data and analyses. All authors contributed to interpretation of the findings and reviewed, commented on, and approved the manuscript before submission for publication.

## Data Availability

All data produced in the present work are contained in the manuscript

## Acknowledgements

The authors want to thank the USAID Demographic and Health Surveys (DHS) programme for making the data available for usage and all participants who contributed to the surveys.

