## Supplementary Information for "The real-world association between male circumcision and risk of HIV infection in sub-Saharan Africa: a household fixed-effects analysis of 279,351 men from 29 countries"

### SUPPORTING INFORMATION

**Supplementary Figure 1. Data availability by country and year.** Surveys were included when self-reported male circumcision status and HIV blood-test result were available for males between 15 years of age and 49 years or older.

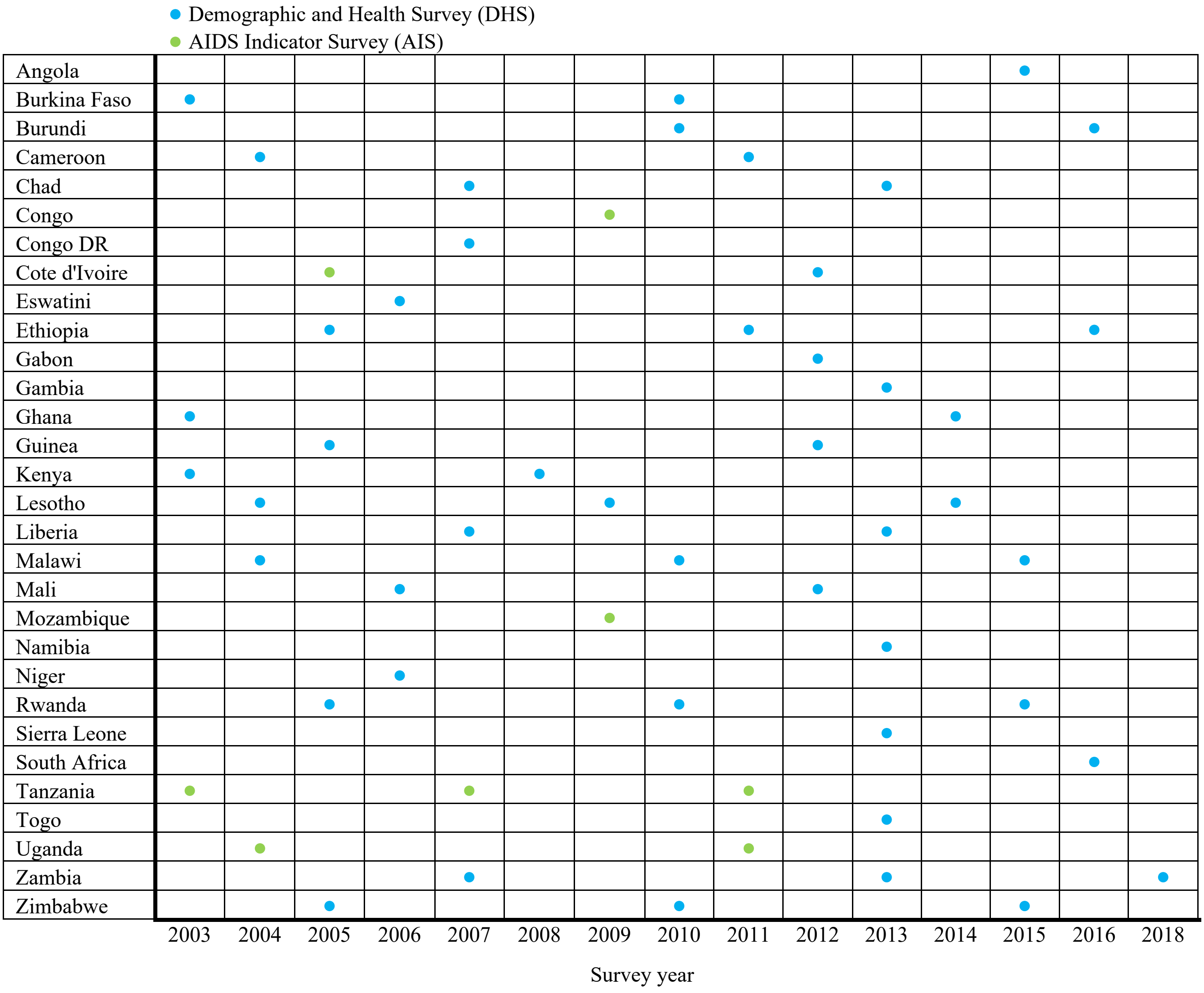

**Supplementary Figure 2. Flow chart of the data selection.**

| | Sub-Saharan Africa, overall | Central Africa | Eastern Africa | Southern Africa | Western Africa |
|---|---|---|---|---|---|
| Available **Demographic and Health Surveys** (DHS) and **AIDS Indicator Surveys** (AIS) conducted in **sub-Saharan Africa** between **2000 and 2020** | **38** countries, **107** surveys | **7 countries, 14 surveys** | **13 countries, 44 surveys** | **4 countries, 8 surveys** | **14 countries, 41 surveys** |
| **Surveys** that include both **HIV and male circumcision indicators** | **31** countries, **61** surveys, **344,832** males | **5** countries, **7** surveys, **41,388** males | **5** countries, **12** surveys, **83,661** males | **9** countries, **18** surveys, **98,768** males | **12** countries, **24** surveys, **121,015** males |
| All men with known **HIV and circumcision status** | **279,351** males | **27,239** males | **63,687** males | **93,821** males | **94,604** males |
| **Two or more adult men per household** | **94,609** males | **10,600** males | **22,306** males | **33,208** males | **28,495** males |
| Two or more adult men per **household** where **at least one men is uncircumcised and one is circumcised** (discordant households) | **16,846** males | **2,763** males | **4,079** males | **7,963** males | **2,041** males |

**Supplementary Table 1. Start year of national voluntary medical male circumcision (VMMC) programmes in Eastern and Southern Africa – for the countries that were included in this study.** Source: WHO and UNAIDS VMMC progress report (https://www.who.int/publications/i/item/voluntary-medical-male-circumcision-progress-brief-2019).

| Country | Start year of national VMMC program |
|---|---|
| eSwatini | 2008 |
| Ethiopia | 2009 |
| Kenya | 2008 |
| Lesotho | 2012 |
| Malawi | 2008 |
| Mozambique | 2009 |
| Namibia | 2009 |
| Rwanda | 2010 |
| South Africa | 2008 |
| South Sudan | 2017 |
| Tanzania | 2009 |
| Uganda | 2010 |
| Zambia | 2008 |
| Zimbabwe | 2009 |

**Supplementary Figure 3. Maps show the African Regions and countries with national voluntary medical male circumcision (VMMC) programs.**

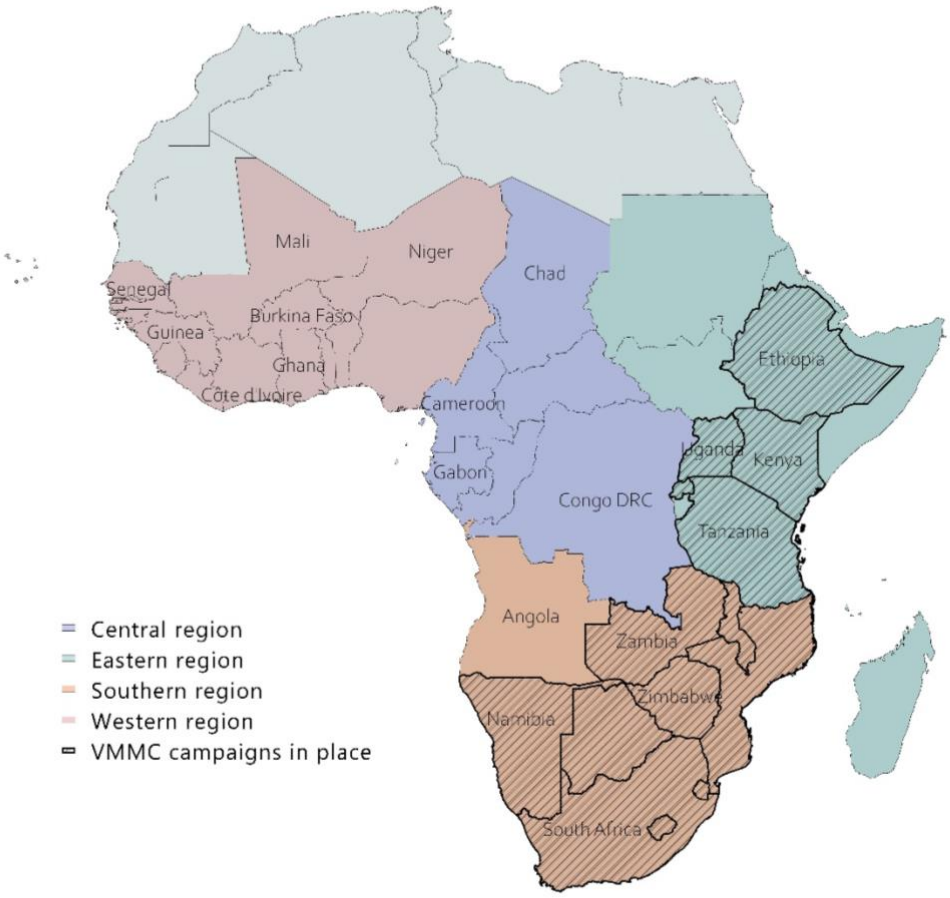

**Supplementary Table 2. Modified Poisson regression models of the effect of male circumcision on HIV status among men in sub-Saharan Africa.** Adjusted risk ratios (aRR) are shown with 95% confidence intervals (CIs), which are adjusted for heterogeneity at the household-level using fixed-effects. Schooling levels are defined as the highest level of education attended. Previously married includes being divorced, separated, or widowed. The number of observations (N) includes men from all households where at least two circumcision discordant men (*i.e.* at least one man circumcised and one uncircumcised) were included in the data. Residual degrees of freedom (DF) indicate the number of independent parameters estimated by the model. The pseudo $R^2$ represents the proportion of the total variability explained by the model.

| | **Model 1 – nested model** | | **Model 2 – adjusted for age** | | **Model 3 – adjusted for age and educational level** | | **Model 4 – adjusted for age, educational level and marital status** | |
|---|---|---|---|---|---|---|---|---|
| **Variable** | **aRR [95% CI]** | **p-value** | **aRR [95% CI]** | **p-value** | **aRR [95% CI]** | **p-value** | **aRR [95% CI]** | **p-value** |
| **Male circumcision status** | | | | | | | | |
| Circumcised | 0.83 [0.76–0.90] | <0.001 | 0.81 [0.74–0.88] | <0.001 | 0.81 [0.74–0.88] | <0.001 | 0.80 [0.73–0.88] | <0.001 |
| Not circumcised | 1 (ref) | - | 1 (ref) | - | 1 (ref) | | 1 (ref) | - |
| **Age** | | | | | | | | |
| 15-24 | | | 1 (ref) | - | 1 (ref) | - | 1 (ref) | - |
| 25-34 | | | 3.32 [3.04–3.63] | <0.001 | 3.32 [3.04–3.64] | <0.001 | 2.57 [3.24–4.12] | <0.001 |
| 35-44 | | | 4.79 [4.39–5.23] | <0.001 | 4.81 [4.40–5.26] | <0.001 | 3.22 [5.13–6.60] | <0.001 |
| 45-54 | | | 5.19 [4.73–5.70] | <0.001 | 5.21 [4.74–5.73] | <0.001 | 3.42 [5.70–7.49] | <0.001 |
| 55+ | | | 3.68 [3.07–4.41] | <0.001 | 3.69 [3.07–4.42] | <0.001 | 2.29 [2.92–4.60] | <0.001 |
| **Highest education** | | | | | | | | |
| No education | | | | | 1.04 [0.92–1.17] | 0.541 | 1.06 [0.87–1.27] | 0.612 |
| Primary | | | | | 1 (ref) | - | 1 (ref) | - |
| Secondary | | | | | 1.03 [0.95–1.12] | 0.448 | 1.03 [0.96–1.18] | 0.256 |
| Post-secondary | | | | | 0.98 [0.84–1.14] | 0.796 | 0.98 [0.77–1.13] | 0.476 |
| **Marital status** | | | | | | | | |
| Never married | | | | | | | 1 | - |

| | | | | | |
|---|---|---|---|---|---|
| Currently married | | | | 1.56 [1.40–1.72] | <0.001 |
| Previously married | | | | 2.13 [1.88–2.40] | <0.001 |
| **Model summary** | | | | | |
| N (observations) | 19,911 | 19,911 | 19,910 | 18,836 | 19,910 |
| Pseudo $R^2$ | 0.169 | 0.233 | 0.233 | 0.239 | 0.237 |

**Supplementary Table 3. Modified Poisson regression model of the estimated effect of male circumcision on living with HIV (males aged 15 years or older) in sub-Saharan Africa, based on the sub-sample of men who live in the same household but differ in male circumcision status (discordant men).**

| | **Model – adjusted for age, educational level, and marital status** | |
|---|---|---|
| **Variable** | **aRR [95% CI]** | **p-value** |
| **Male circumcision status** | | |
| Circumcised | 0.80 [0.73–0.88] | <0.001 |
| Not circumcised | 1 (ref) | - |
| **Age** | | |
| 15-24 | 1 (ref) | - |
| 25-34 | 2.34 [1.92–2.86] | <0.001 |
| 35-44 | 3.01 [2.39–3.79] | <0.001 |
| 45-54 | 3.19 [2.50–4.06] | <0.001 |
| 55+ | 2.01 [1.40–2.88] | <0.001 |
| **Highest education** | | |
| No education | 1.05 [0.85–1.30] | 0.651 |
| Primary | 1 (ref) | - |
| Secondary | 0.96 [0.83–1.10] | 0.536 |
| Post-secondary | 1.07 [0.82–1.40] | 0.594 |
| **Marital status** | | |
| Never married | 1 | - |
| Currently married | 1.53 [1.26–1.86] | <0.001 |
| Previously married | 1.86 [1.48–2.33] | <0.001 |
| **Model summary** | | |
| N (observations) | 7,759 | |
| Residual DF | 6,669 | |
| Pseudo $R^2$ | 0.277 | |

**Supplementary Table 4. Outcomes for sub-group analyses.** Adjusted risk ratios (aRR) are shown with 95% confidence intervals (CIs), which are adjusted for heterogeneity at the household-level using fixed-effects. The number of observations (N) includes men from all households where at least two circumcision discordant men (*i.e.* at least one man circumcised and one uncircumcised) were included in the data. Residual degrees of freedom (DF) indicate the number of independent parameters estimated by the model. The pseudo $R^2$ represents the proportion of the total variability explained by the model.

| | **By African region** | | | | **By age** | | **By presence national VMMC programs** | | **By family relatedness** |
|---|---|---|---|---|---|---|---|---|---|
| | **Central** | **Eastern** | **Southern** | **Western** | **34 or younger** | **35 or older** | **National VMMC programmes launched** | **No national VMMC programmes launched** | **Brothers** |
| **Model estimates** | | | | | | | | | |
| **Risk ratio** | 0.88 | 0.63 | 0.81 | 0.84 | 0.79 | 0.82 | 0.75 | 0.98 | 0.78 |
| 95% CI | 0.43–1.82 | 0.46–0.85 | 0.72–0.91 | 0.60–1.16 | 0.69–0.91 | 0.63–1.06 | 0.66–0.85 | 0.83–1.15 | 0.66–0.91 |
| Robust SE | 0.33 | 0.10 | 0.05 | 0.14 | 0.057 | 0.110 | 0.046 | 0.083 | 0.064 |
| z-score | -0.35 | -3.04 | -3.60 | -1.07 | -3.28 | -1.51 | -4.69 | -0.29 | -3.07 |
| p-value | 0.727 | 0.002 | <0.001 | 0.285 | <0.001 | 0.130 | <0.001 | 0.768 | 0.002 |
| **Model summary** | | | | | | | | | |
| N (observations) | 419 | 1,235 | 7,021 | 11,235 | 10,247 | 2,778 | 5,868 | 14,043 | 3,622 |
| Residual df | 251 | 729 | 4,065 | 10,663 | 8,590 | 2,415 | 3,410 | 12,329 | 2,111 |
| Pseudo-$R^2$ | 0.168 | 0.159 | 0.158 | 0.170 | 0.208 | 0.176 | 0.151 | 0.230 | 0.100 |

**Supplementary Table 5. Comparison of data characteristics for all men included in the surveys versus men from circumcision discordant households.**

| | All men N (%) | More than one man, discordant circumcision status N (%) |
|---|---|---|
| **Sample size** | 344,832 | 18,182 |
| **HIV status** | | |
| Positive | 15,952 (4.7%) | 1,078 (6.0%) |
| Negative | 322,794 (95.3%) | 16,855 (94.0%) |
| **Male circumcision status** | | |
| Circumcised | 183,460 (64.7%) | 9,184 (53.8%) |
| Not circumcised | 100,088 (35.3%) | 7,904 (46.3%) |
| **Sociodemographic characteristics** | | |
| **Age (per 10-year age group)** | | |
| 34 or younger | 224,050 (65.0%) | 13,065 (71.9%) |
| 35 and older | 120,782 (35.0%) | 5,117 (28.1%) |
| **Highest education** | | |
| No education | 75,403 (21.9%) | 2,117 (11.6%) |
| Primary | 129,201 (37.5%) | 7,138 (39.3%) |
| Secondary | 120,958 (35.1%) | 7,673 (42.2%) |
| Post-secondary | 19,245 (5.6%) | 1,253 (6.9%) |
| **Socioeconomic status** | | |
| 1 'Poorest' | 66,980 (19.4%) | 2,737 (15.1%) |
| 2 | 66,485 (19.3%) | 2,935 (16.1%) |
| 3 | 65,708 (19.1%) | 3,159 (17.4%) |
| 4 | 67,275 (19.5%) | 3,714 (20.4%) |
| 5 'Wealthiest' | 78,381 (22.7%) | 5,637 (31.0%) |
| **Marital status** | | |
| Never married | 141,046 (41.6%) | 10,771 (59.2%) |
| Currently married | 184,183 (54.3%) | 6,639 (36.5%) |
| Previously married | 13,941 (4.1%) | 772 (4.3%) |
| **(Sexual) behavioural characteristics** | | |
| **Lifetime sex partners** | | |
| None | 97,150 (29.2%) | 5,964 (33.7%) |
| 1-2 | 89,459 (26.9%) | 4,166 (23.6%) |
| 3-6 | 88,331 (26.5%) | 4,373 (24.7%) |
| 7+ | 58,127 (17.5%) | 3,179 (18.0%) |
| **Had STI during past 12 months** | | |
| Yes | 18,541 (5.4%) | 1,072 (5.9%) |
| No | 325,962 (94.6%) | 17,096 (94.1%) |
| **Ever engaged in transactional sex** | | |
| Yes | 19,794 (11.1%) | 994 (12.3%) |
| No | 159,146 (88.9%) | 7,066 (87.7%) |
| **Survey characteristics** | | |
| **Region** | | |
| Central Africa | 41,388 (12.0%) | 3,689 (20.3%) |
| Eastern Africa | 83,661 (24.3%) | 4,147 (22.8%) |
| Southern Africa | 98,768 (28.6%) | 8,197 (45.1%) |
| Western Africa | 121,015 (35.1%) | 2,149 (11.8%) |
| **Time of survey** | | |
| Between 2000-2004 | 40,308 (11.7%) | 1,390 (7.6%) |
| Between 2005-2009 | 106,046 (30.8%) | 5,537 (30.45%) |
| Between 2010-2014 | 136,072 (39.5%) | 6,068 (33.4%) |
| Between 2015-2019 | 62,406 (18.1%) | 5,187 (28.5%) |

**Supplementary Table 6. Comparison of data characteristics for circumcised men from single-men households, circumcised men from multiple-men households, uncircumcised men from single-men households, and uncircumcised men from multiple-men households.**

| | Circumcised men from single-men households N (%) | Circumcised men from multiple-men households N (%) | Uncircumcised men from single-men households N (%) | Uncircumcised men from multiple-men households N (%) |
|---|---|---|---|---|
| **Sample size** | 122,532 | 59,164 | 65,038 | 36,814 |
| **HIV status** | | | | |
| Positive | 116,614 (3.6%) | 1,462 (2.5%) | 6,366 (10.0%) | 2,459 (6.8%) |
| Negative | 4,410 (96.4%) | 57,016 (97.5%) | 57,352 (90.0%) | 33,672 (93.2%) |
| **Sociodemographic characteristics** | | | | |
| **Age (per 10-year age group)** | | | | |
| 34 or younger | 73,801 (60.2%) | 42,644 (72.1%) | 40,943 (63.0%) | 26,801 (72.8%) |
| 35 and older | 48,731 (39.8%) | 16,520 (27.9%) | 24,095 (37.0%) | 10,013 (27.2%) |
| **Highest education** | | | | |
| No education | 36,723 (30.0%) | 12,016 (20.3%) | 7,720 (11.9%) | 3,043 (8.3%) |
| Primary | 39,270 (32.1%) | 19,569 (33.1%) | 31,515 (48.5%) | 17,414 (47.3%) |
| Secondary | 38,887 (31.7%) | 23,656 (40.0%) | 22,563 (34.7%) | 14,704 (39.9%) |
| Post-secondary | 7,647 (6.2%) | 3,920 (6.6%) | 3,235 (5.0%) | 1,652 (4.5%) |
| **Socioeconomic status** | | | | |
| 1 'Poorest' | 25,748 (21.0%) | 8,982 (15.2%) | 13,919 (21.4%) | 5,453 (14.8%) |
| 2 | 24,519 (20.0%) | 9,764 (16.5%) | 13,444 (20.7%) | 6,462 (17.6%) |
| 3 | 22,748 (18.6%) | 10,583 (17.9%) | 13,035 (20.4%) | 7,636 (20.7%) |
| 4 | 22,084 (18.0%) | 12,239 (20.7%) | 13,008 (20.0%) | 8,179 (22.2%) |
| 5 'Wealthiest' | 27,432 (22.4%) | 17,596 (29.4%) | 11,632 (17.9%) | 9,082 (24.7%) |
| **Marital status** | | | | |
| Never married | 78,995 (64.5%) | 21,469 (36.3%) | 41,757 (64.2%) | 12,172 (34.5%) |
| Currently married | 38,031 (31.0%) | 35,359 (59.8%) | 20,407 (31.4%) | 23,344 (63.4%) |
| Previously married | 5,506 (4.5%) | 2,335 (4.0%) | 2,872 (4.4%) | 1,298 (3.5%) |
| **(Sexual) behavioural characteristics** | | | | |
| **Lifetime sex partners** | | | | |
| None | 27,562 (23.7%) | 23,156 (40.7%) | 12,916 (20.3%) | 13,742 (38.0%) |
| 1-2 | 31,506 (27.0%) | 12,375 (21.7%) | 20,840 (32.8%) | 9,505 (26.3%) |
| 3-6 | 32,508 (27.9%) | 12,331 (21.7%) | 20,417 (32.1%) | 8,898 (24.6%) |
| 7+ | 24,988 (21.4%) | 9,089 (16.0%) | 9,443 (14.8%) | 4,056 (11.2%) |
| **Had STI during past 12 months** | | | | |
| Yes | 6,969 (5.7%) | 2,893 (4.9%) | 4,033 (6.2%) | 1,801 (4.9%) |
| No | 115,472 (94.3%) | 56,233 (95.1%) | 60,951 (93.8%) | 34,976 (95.1%) |
| **Ever engaged in transactional sex** | | | | |
| Yes | 6,497 (9.9%) | 2,895 (10.2%) | 4,558 (12.2%) | 2,028 (12.4%) |
| No | 59,264 (90.1%) | 25,640 (89.9%) | 32,912 (87.8%) | 14,280 (87.6%) |
| **Survey characteristics** | | | | |
| **Region** | | | | |
| Central Africa | 12,192 (10.0%) | 7,933 (13.4%) | 4,459 (6.9%) | 2,683 (7.3%) |
| Eastern Africa | 25,896 (21.1%) | 13,351 (22.6%) | 15,943 (24.5%) | 9,194 (25.0%) |
| Southern Africa | 20,340 (16.6%) | 10,423 (17.6%) | 41,855 (64.4%) | 23,555 (64.0%) |
| Western Africa | 64,104 (52.3%) | 27,457 (46.4%) | 2,781 (4.3%) | 1,382 (3.8%) |
| **Time of survey** | | | | |
| Between 2000-2004 | 18,891 (15.4%) | 9,962 (16.8%) | 2,416 (3.7%) | 1,499 (4.1%) |
| Between 2005-2009 | 50,256 (41.0%) | 19,231 (32.5%) | 15,420 (23.7%) | 8,449 (23.0%) |
| Between 2010-2014 | 38,568 (31.5%) | 21,790 (36.3%) | 28,368 (43.6%) | 16,578 (45.0%) |
| Between 2015-2019 | 14,817 (12.1%) | 8,181 (13.8%) | 18,834 (29.0%) | 10,288 (28.0%) |
